# MRI-Derived Patterns of Post-Cardiac Arrest Brain Injury: Linking Anatomical Injury and Cerebrovascular Physiology

**DOI:** 10.64898/2026.09.03.26362123

**Authors:** Connor A. Larkey, Gaurav Ambwani, Warda Limaye, Sophie Stukas, Jennifer Cooper, Cheryl L. Wellington, Peter A. Gooderham, Denise Foster, Donald E. Griesdale, Mypinder S. Sekhon, William Guest, Ryan L. Hoiland

**Affiliations:** Department of Cellular and Physiological Sciences, Faculty of Medicine, University of British Columbia, Vancouver, British Columbia, Canada; Djavad Mowafaghian Centre for Brain Health, University of British Columbia, Vancouver, British Columbia, Canada; Centre for Chronic Disease Prevention and Management, University of British Columbia, Kelowna, British Columbia; Department of Radiology, Faculty of Medicine, University of British Columbia, Vancouver, British Columbia, Canada; Department of Pathology and Laboratory Medicine, Faculty of Medicine, University of British Columbia, Vancouver, British Columbia, Canada; Division of Neurosurgery, Department of Surgery, Vancouver General Hospital, University of British Columbia, Vancouver, BC, Canada; Division of Critical Care Medicine, Department of Medicine, Vancouver General Hospital, University of British Columbia; Department of Anesthesiology, Pharmacology, and Therapeutics, University of British Columbia, Vancouver, British Columbia, Canada; Centre for Clinical Epidemiology and Evaluation, Vancouver Coastal Health Research Institute, University of British Columbia, Vancouver, Canada; International Collaboration on Repair Discoveries, University of British Columbia, Vancouver, British Columbia, Canada

**Keywords:** Cardiac arrest, Post-cardiac arrest brain injury, Brain tissue oxygen tension (PbtO_2_), Magnetic resonance imaging (MRI), Neurologic biomarkers, Cerebrovascular physiology

## Abstract

**Introduction:** Post-cardiac arrest brain injury (PCABI) patterns across patients are heterogeneous, and conventional clinical variables incompletely capture in vivo injury severity. Here, we evaluated whether anatomically resolved diffusion-weighted MRI (DWI) lesion patterns identify biologically distinct PCABI phenotypes.

**Methods:** We conducted a retrospective study of patients with PCABI who underwent brain MRI and intraparenchymal brain tissue oxygen tension (PbtO_2_) monitoring (n = 24). Ischemic lesion burden was quantified using an MNI atlas-based apparent diffusion coefficient pipeline with a voxel intensity threshold of <650×10^-6^ mm²/s and was summarized at whole-brain, hemispheric, tissue-class, and segment-levels. Principal component analysis (PCA) and k-means clustering were used to identify MRI-derived injury patterns for downstream physiological analysis.

**Results:** Greater whole-brain lesion burden was associated with lower mean PbtO_2_ (r = -0.49, p = 0.014), with concurrent associations in the left hemisphere (r = -0.50, p = 0.012), right hemisphere (r = -0.48, p = 0.019), grey matter (r = -0.48, p = 0.018), and white matter (r = -0.49, p = 0.016). PCA identified a dominant lesion-pattern axis associated with PbtO_2_ (r = 0.48, p = 0.0169). K-means clustering in PC1- PC2 space identified three MRI-derived injury patterns with distinct regional signatures and progressively greater whole-brain lesion burden (p < 0.001).

**Discussion:** Greater MRI-defined lesion burden was associated with lower PbtO_2_ post-cardiac arrest. Anatomically resolved DWI/ADC lesion mapping identified three MRI-derived injury patterns that differed in cerebrovascular physiology, lesion patterning and whole-brain injury burden. Integrating quantitative MRI, intraparenchymal neuromonitoring, and blood biomarkers improved the characterization of the heterogeneity observed in PCABI patients.

**HIGHLIGHTS:**

- Atlas DWI/ADC mapping identified three MRI-derived injury patterns.
- MRI-derived patterns captured regional injury beyond whole-brain burden.
- Greater ischemic lesion burden was associated with lower PbtO_2_.
- MRI-derived patterns differed in PbtO_2_ and selected biomarkers.
- Brain injury patterns were not explained by admission physiology nor time to ROSC.

## INTRODUCTION

Post-cardiac arrest brain injury (PCABI) remains the most prevalent cause of death and disability after successful resuscitation.^1–3^ Historically viewed as a diffuse hypoxic-ischemic insult, PCABI is now recognized as a dynamic post-resuscitation syndrome involving ischemic depolarization, reperfusion injury, dysregulated cerebral perfusion, microvascular failure, impaired oxygen diffusion, neuroinflammation, edema, and disordered cellular oxygen use.^1,2,4,5^ These complexities have contributed to the neutral or mixed results of trials targeting isolated interventions for systemic variables, such as temperature, oxygenation, blood pressure, and carbon dioxide targets.^6–12^ Current frameworks therefore emphasize phenotyping PCABI according to *in vivo* injury severity and mechanism, rather than purely inferring injury biology from historical arrest characteristics.^13–18^

Recent efforts to phenotype PCABI have used clinical, electrophysiologic, neuroimaging, and post-ROSC cerebrovascular approaches.^15–28^ For example, phenotypes have been described pertaining to brain tissue oxygen tension (PbtO_2_),^21,22,24,27^ intracranial pressure,^22,26^ jugular venous oxygen saturation,^23,25^ and limitations for oxygen delivery and extraction.^24,25,27,28^ Low PbtO_2_ (<20 mmHg) in patients with PCABI has been associated with ongoing neuroglial injury,^21^ highlighting the importance of understanding the cerebrovascular physiology and its implications for disease severity and progression.^13,14,27^

While it is clear that PCABI can manifest distinct cerebrovascular changes across patients, how post-ROSC cerebrovascular physiology is influenced by brain injury patterns remains unexplored. Diffusion-weighted imaging (DWI) and apparent diffusion coefficient (ADC) maps are among the most established tools for quantifying PCABI.^29–37^ Restricted diffusion reflects cytotoxic edema, evolves over time, and has repeatedly demonstrated prognostic value when assessed quantitatively in the early post-arrest period.^29–32,35–44^ However, conventional quantitative MRI approaches often reduce injury to mean ADC, whole-brain lesion burden, or specific regions of interest.^29–32,37–43^ This is limiting because post-arrest diffusion abnormalities are anatomically heterogeneous and may show spatiotemporal-specific prognostic patterns.^30,33–35,44–47^

We previously developed a semi-automated atlas-based DWI/ADC pipeline to quantify ischemic lesion burden with segment-level anatomical resolution in PCABI.^48^ Building on this work, we integrated anatomically resolved DWI/ADC lesion quantification with intraparenchymal PbtO_2_ monitoring and paired arterial-jugular venous differences of blood-based brain injury biomarkers in patients with PCABI. We examined whether global MRI-defined lesion burden is associated with changes in PbtO_2_ and whether certain MRI-derived injury patterns identified through unsupervised dimensionality reduction of regional lesion features reflect unique post-ROSC cerebrovascular physiology. We hypothesized that regional DWI/ADC lesion patterning would capture physiologically meaningful heterogeneity in PCABI.

## METHODS

### Study Design

We conducted a retrospective study of patients with PCABI admitted to the intensive care unit (ICU) at Vancouver General Hospital. The study received ethical approval from the University of British Columbia Clinical Research Ethics Board (H24-00530) and operational approval from Vancouver Coastal Health Research Institute (V24-00530). The requirement for individual participant consent was waived as part of the approved retrospective records-review protocol. Patients were eligible if they had a diagnosis of PCABI, clinically indicated brain MRI including DWI, ADC, and susceptibility-weighted imaging (SWI) sequences, and available intraparenchymal PbtO_2_ monitoring data. Patients were excluded if SWI demonstrated localized hemorrhage signal confounding ADC-defined lesion quantification. Patients with concurrent traumatic brain injury were also excluded. Clinical and demographic variables were extracted from the British Columbia Critical Care Database, electronic medical records, and imaging records.

### Intraparenchymal PbtO_2_ monitoring

Patients underwent intraparenchymal PbtO_2_ monitoring using a Licox catheter (Integra LifeSciences, Plainsboro, NJ, USA) inserted through a cranial access bolt in the right, non-dominant frontal region (Figure 1A). The PbtO_2_ probe was positioned in frontal subcortical white matter, and placement was confirmed with CT imaging. PbtO_2_ data were recorded using ICM+ brain monitoring software (Cambridge Enterprise, Cambridge, United Kingdom). For the present analysis, PbtO_2_ was summarized as the mean value over the first 24 hours of monitoring. For patients who died before 24 hours of monitoring were complete, mean PbtO_2_ was calculated using all available data before death.

**Figure 1.**
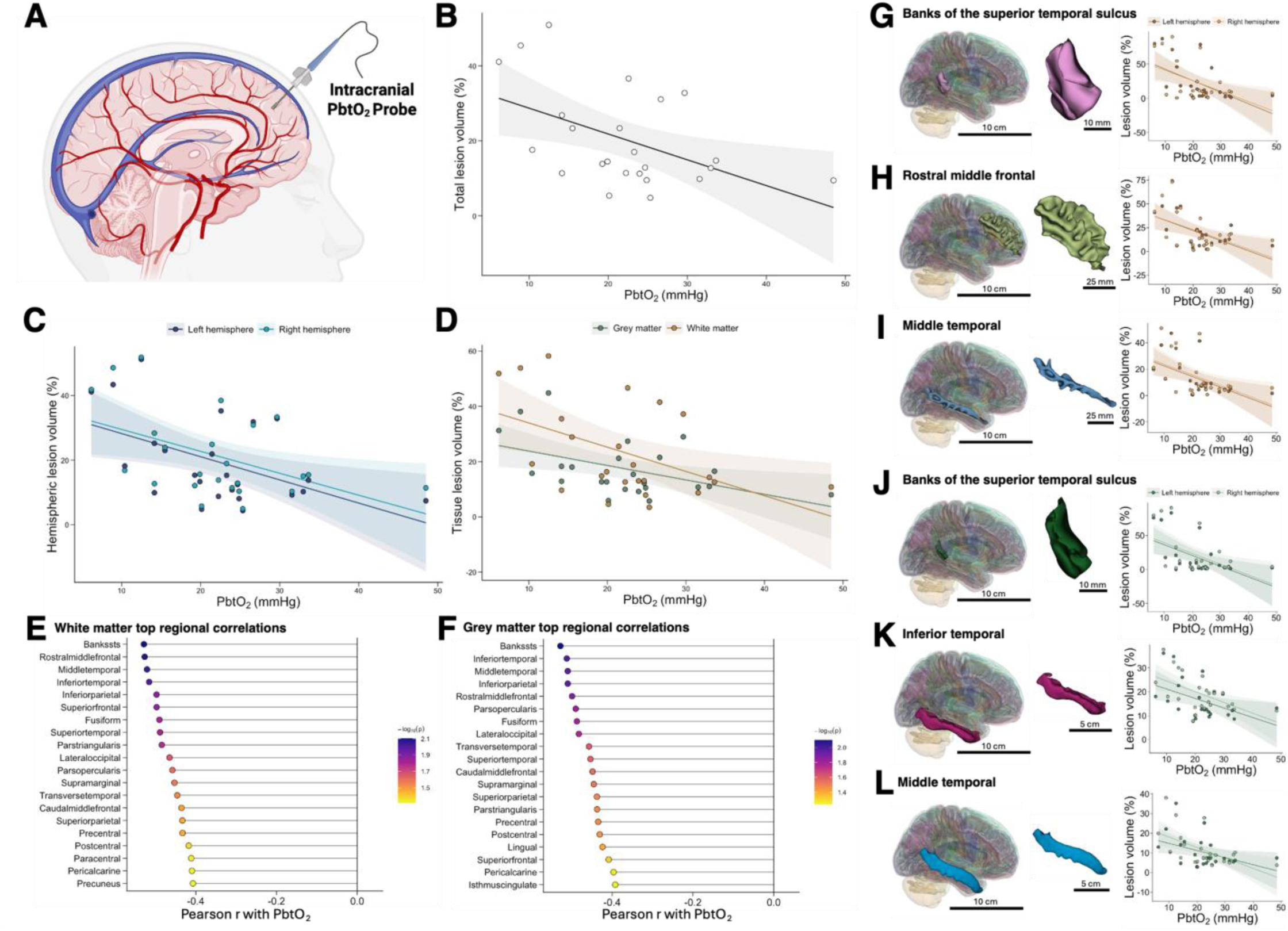
Associations between MRI-defined ischemic lesion burden and brain tissue oxygen tension (PbtO_2_) after cardiac arrest. (A) Schematic illustrating typical intraparenchymal brain tissue oxygen tension (PbtO_2_) probe placement in patients with post-cardiac arrest brain injury. (B) Whole-brain lesion burden was inversely associated with mean PbtO_2_ (r = -0.49, p = 0.014; n = 24). (C) Hemisphere-specific lesion burden was inversely associated with PbtO_2_ in the left hemisphere (r = -0.50, p = 0.012) and right hemisphere (r = -0.48, p = 0.019). (D) Grey matter and white matter lesion burden were both inversely associated with PbtO_2_ (grey matter: r = -0.48, p = 0.018; white matter: r = -0.49, p = 0.016). (E-F) Top 20 bilateral white matter and grey matter regions ranked by the strength of association between regional lesion burden and PbtO_2_. (G-I) The three white matter regions with the strongest inverse associations were banks of the superior temporal sulcus (r = -0.53, p = 0.008), rostral middle frontal region (r = -0.53, p = 0.008), and middle temporal region (r = -0.52, p = 0.009). (J-L) The three grey matter regions with the strongest inverse associations were banks of the superior temporal sulcus (r = -0.53, p = 0.008), inferior temporal region (r = -0.51, p = 0.010), and middle temporal region (r = -0.51, p = 0.011). Anatomical renderings were generated in 3D Slicer and corresponding correlation plots were generated in R.

### MRI acquisition and regional lesion analyses

MRI was acquired on 1.5 or 3 Tesla Siemens MRI scanners using standard non-contrast brain imaging sequences and performed as clinically indicated. At minimum, imaging included axial DWI, corresponding ADC maps, SWI, and T2 fluid-attenuated inversion recovery sequences.

MRI lesion burden was quantified using our previously described semi-automated DWI/ADC pipeline.^48^ DWI and SWI were preprocessed in 3D Slicer, skull stripped using HD-BET, and registered to the MNI152 1-mm template using Elastix.^49–52^ FreeSurfer *aparc*+*aseg* and *wmparc* parcellations were overlaid on registered ADC maps.^53,54^ Ischemic injury was defined as ADC <650 × 10^-6^ mm^2^/s, and lesion burden was calculated as the percentage of voxels below threshold within each segment.^37,39–43^ Lesion percentages were extracted by hemisphere and tissue class after exclusion of non-parenchymal/non-informative labels, yielding 164 segments per patient.

Associations between mean PbtO_2_ and whole-brain, hemispheric, and tissue class lesion burden were assessed using Pearson correlations. Bilateral regional rankings of Pearson correlation r values for relative lesion burden of all segments were generated by averaging left and right hemisphere lesion percentages for each segment within each tissue class.

### MRI lesion patterns

MRI-derived injury patterns were classified using dimensionality reduction and unsupervised clustering based only on anatomical lesion burden. Segment-level lesion percentages were arranged into a feature-by-patient matrix, with each feature defined by anatomical segment, tissue class, and hemisphere. Each MRI lesion feature was z-scored prior to principal component analysis (PCA).

PCA was performed on the retained feature matrix. K-means clustering (k=3) was then applied to PC1 and PC2 using 100 random starts, and a fixed random seed for reproducibility analysis. Clinical, physiological, biomarker, and outcome variables were not included in PCA, k-means clustering, or feature selection, and were only joined after cluster assignment for downstream PCABI characterization. The k = 3 solution was evaluated using within-cluster sum of squares, average silhouette width, and repeated-seed stability. MRI features contributing most to cluster separation were identified by ranking Kruskal-Wallis tests across clusters and pairwise Wilcoxon tests for cluster comparison. Heatmaps were generated from z-scored lesion percentages.

### Blood-Based Neurologic Biomarkers

Analyses were performed using previously described methods.^21^ Simultaneous arterial and jugular venous blood samples were collected into serum separator tubes during the first 24 hours of neuromonitoring (Figure 4A). All blood samples included in the present analysis were collected prior to MRI, which was performed subsequently at a median of 3.0 [1.0, 8.0] days post-arrest. Samples were set upright in the dark for 10 minutes and then centrifuged at 600g for 10 minutes. Serum supernatant was aliquoted into cryovials and immediately stored at -80°C until analysis.

**Figure 2.**
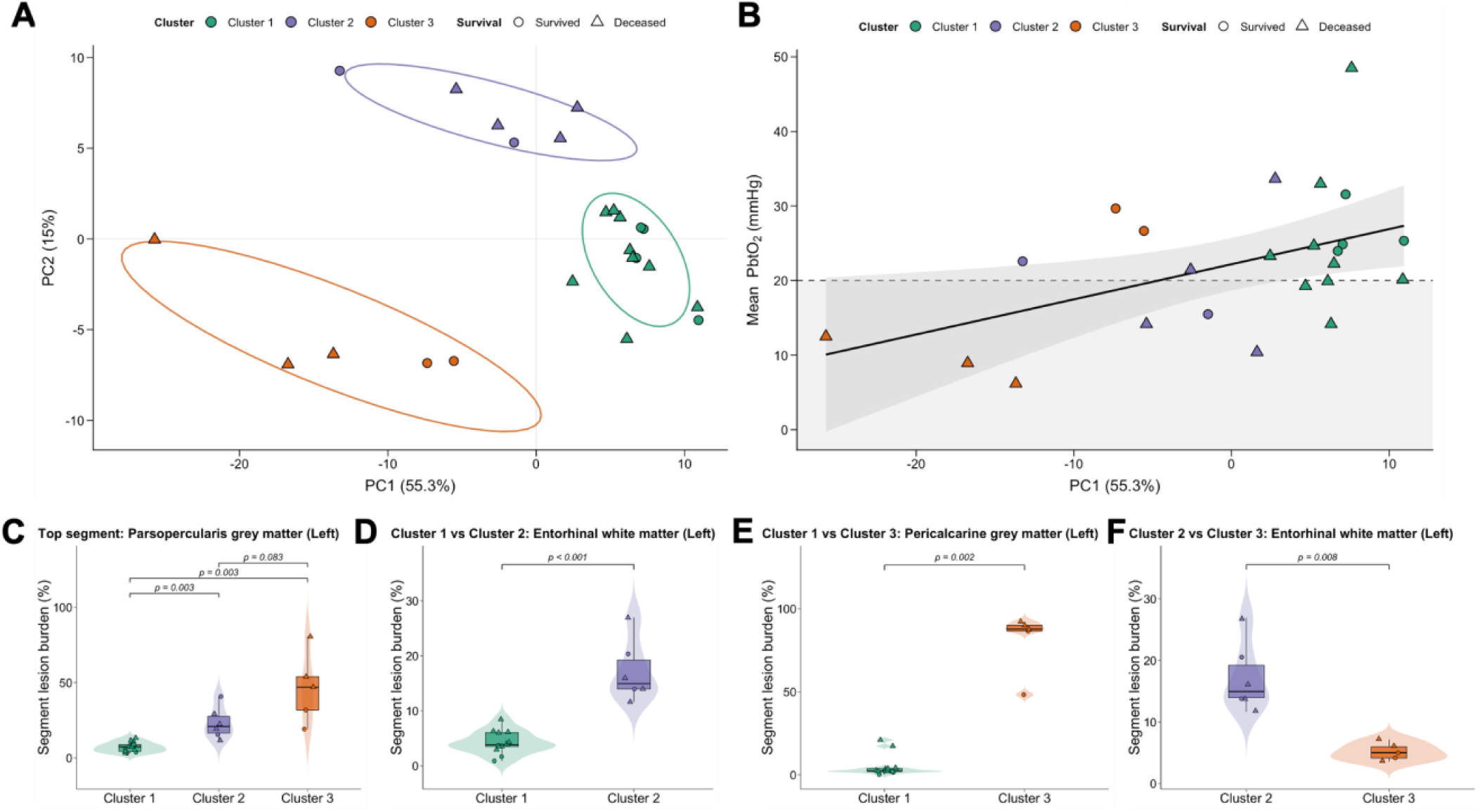
MRI-derived clustering based on regional DWI/ADC lesion patterns. (A) Principal component analysis (PCA) of anatomically resolved MRI lesion features. Each point represents one patient (n = 24), coloured by k-means cluster assignment in PC1-PC2 space (k = 3); circles indicate survivors and triangles indicate deceased patients. Ellipses represent 68% confidence ellipses around each cluster. (B) PC1 scores were associated with mean brain tissue oxygen tension (PbtO_2_; Pearson r = 0.48, p = 0.0169). The dashed horizontal line indicates PbtO_2_ = 20 mmHg. The shaded band represents the 95% confidence interval around the fitted regression line. (C-F) Regional lesion burden for MRI features that most strongly discriminated cluster membership. Panel C shows the top overall discriminatory feature across all clusters by Kruskal-Wallis testing (p < 0.001). Panels D-F show the top pairwise discriminatory features for Cluster 1 versus Cluster 2, Cluster 1 versus Cluster 3, and Cluster 2 versus Cluster 3, respectively, with Holm-adjusted pairwise Wilcoxon rank-sum p-values shown. Values are regional lesion burden percentages. Violin plots show the distribution of individual patient values; embedded boxplots show the median and interquartile range, with whiskers extending to 1.5 × interquartile range. K-means clustering was performed using PC1 and PC2 derived from PCA of regional MRI lesion features alone; clinical, physiological, biomarker, and outcome variables were not included in cluster assignment. Cluster labels are nominal and were assigned after unsupervised clustering. Cluster 1 included 13 patients, Cluster 2 included 6 patients, and Cluster 3 included 5 patients.

**Figure 3.**
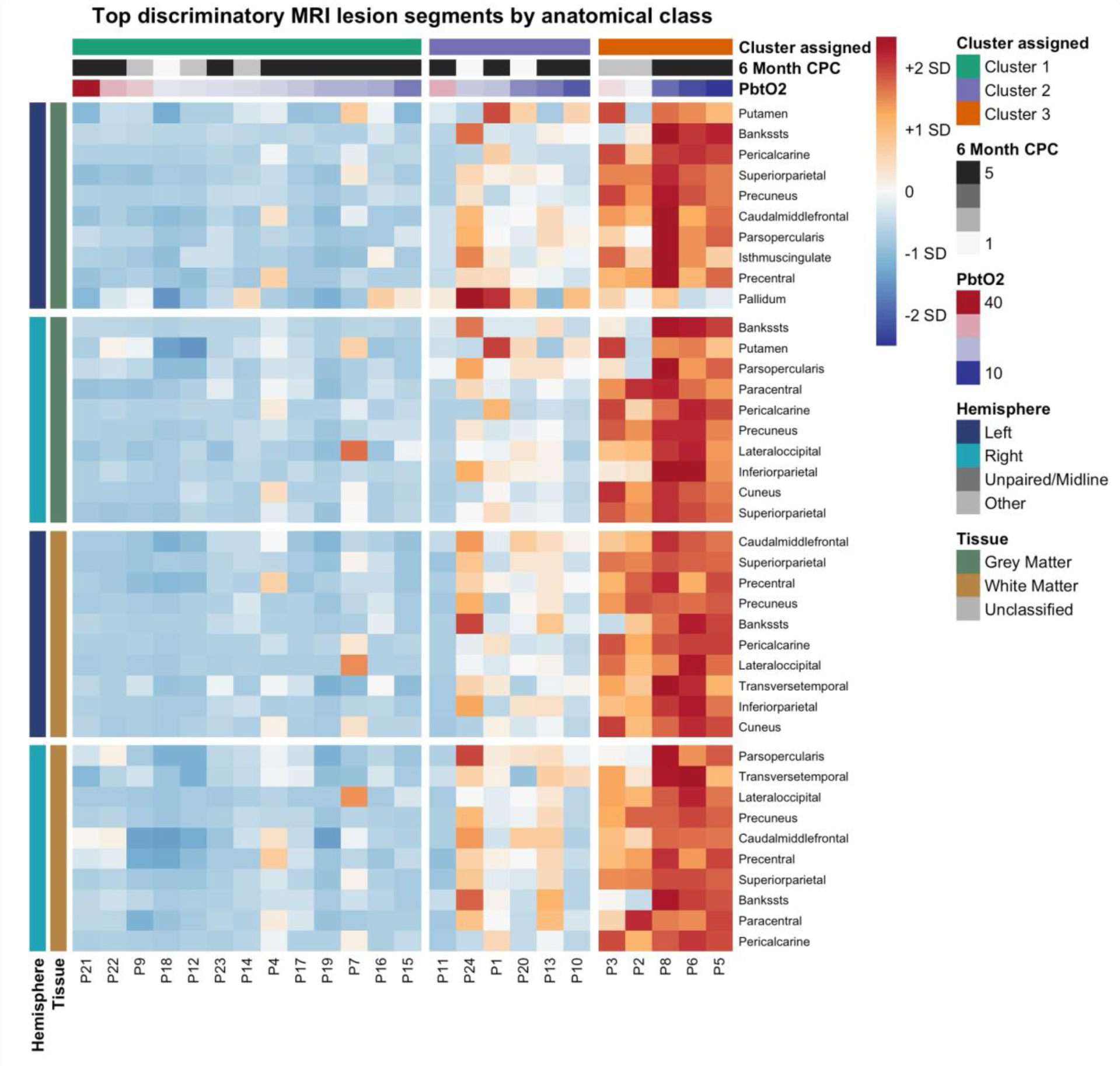
MRI lesion-pattern heatmap across MRI-derived clusters. Heatmap of standardized MRI lesion feature z-scores across patients grouped by cluster assignment. Columns represent individual patients, and rows represent the top 10 cluster-discriminatory lesion features selected within each anatomical category: left grey matter, right grey matter, left white matter, and right white matter. Patients are ordered by cluster assignment and, within each cluster, by mean brain tissue oxygen tension (PbtO_2_). Annotation tracks display cluster membership, 6-month Cerebral Performance Category (CPC), and mean PbtO_2_.

**Figure 4.**
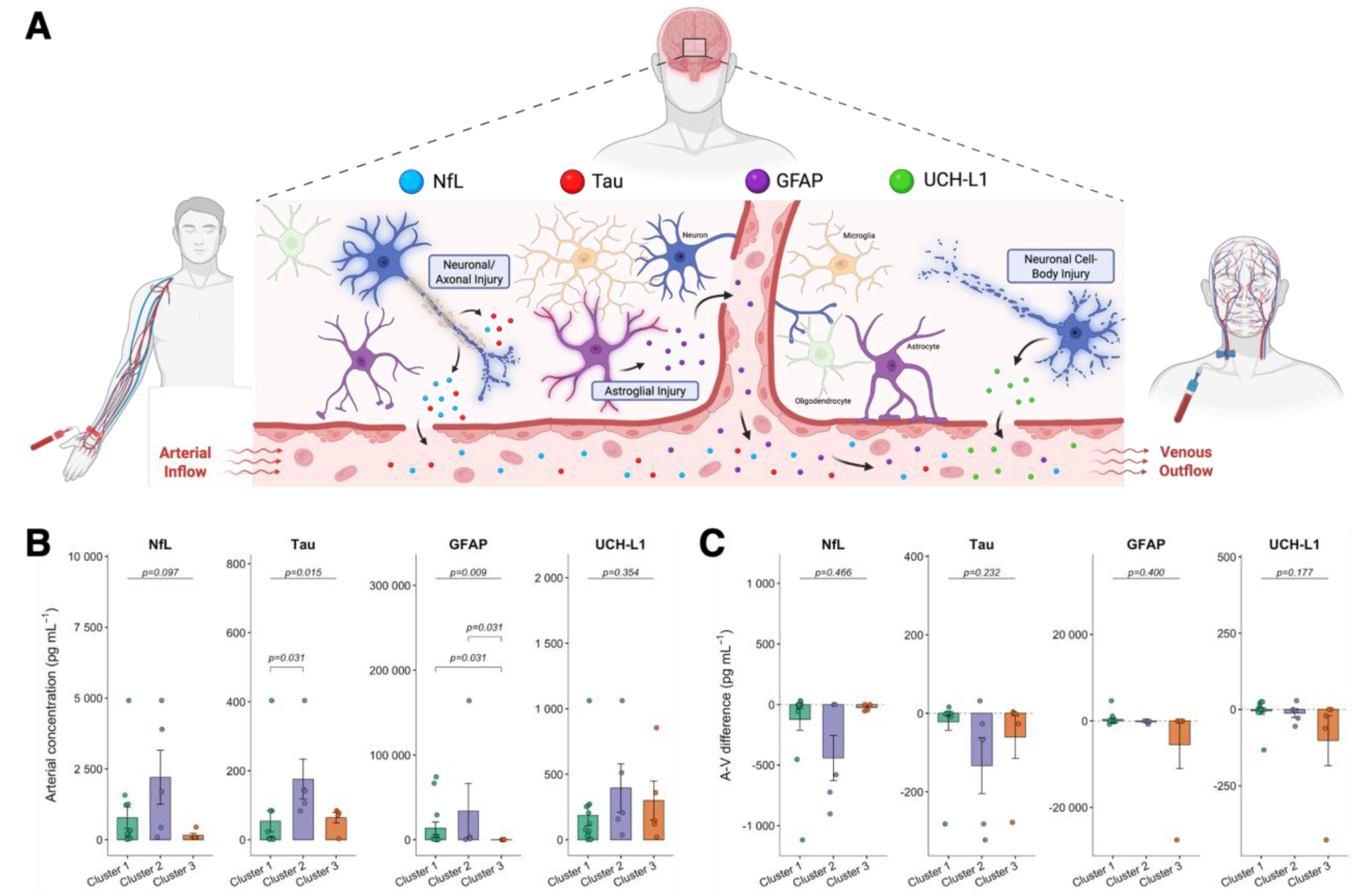
Blood-based neurologic biomarkers across MRI-derived injury patterns. Paired arterial and jugular venous blood samples were examined to evaluate circulating neurologic biomarkers and arterial-venous concentration differences across MRI-derived injury patterns. (A) Schematic illustrating radial arterial and jugular bulb venous blood sampling following post-cardiac arrest brain injury (PCABI). Neurofilament light chain (NfL), total tau, glial fibrillary acidic protein (GFAP), and ubiquitin C-terminal hydrolase L1 (UCH-L1) were measured in arterial and jugular venous serum. (B) Arterial concentrations of NfL, tau, GFAP, and UCH-L1 stratified by MRI-derived injury pattern. Arterial tau concentrations differed between Clusters 1 and 2. Arterial GFAP concentrations differed between Clusters 1 and 3 and between Clusters 2 and 3. (C) Arterial-venous concentration differences (av_DIFF_) for NfL, tau, GFAP, and UCH-L1 stratified by MRI-derived injury pattern. Negative av_DIFF_ values indicate higher jugular venous than arterial biomarker concentrations. Bars represent median values, error bars represent the interquartile range, and points represent individual patients. Biomarker-wise group differences were assessed using Kruskal-Wallis tests, with p-values shown above each biomarker panel. Holm-adjusted pairwise Wilcoxon rank-sum tests are shown where significant.

Serum samples were analyzed using the Quanterix single molecule array (Simoa) platform with the HD-1 analyzer. Biomarkers in the present analysis included neurofilament light chain (NfL), total tau, glial fibrillary acidic protein (GFAP), and ubiquitin carboxy terminal hydrolase L1 (UCH-L1). Arterial concentrations were analyzed as systemic circulating biomarker measures. Arterial-to-jugular venous differences (av_DIFF_) were calculated as arterial concentration minus jugular venous concentration, such that negative values indicated higher jugular venous than arterial concentrations. To account for differences that may arise simply due to measurement variability, calculated av_DIFF_ needed to be greater than the absolute coefficient of variation in order to be included in the dataset. Values for av_DIFF_ that did not meet this criterion were considered zero.

### Admission physiology and ROSC analyses

Admission arterial pH, lactate, serum creatinine, and PaCO_2_ were compared across MRI-derived clusters using Kruskal Wallis tests. Associations between time to ROSC, mean PbtO_2_, and whole-brain lesion burden were assessed via Pearson correlation.

### Statistical analysis

Continuous variables are summarized as median [interquartile range], unless otherwise specified, and categorical variables as counts and percentages. All correlations were assessed via Pearson correlations. All differences between MRI-derived clusters were first assessed using Kruskal- Wallis tests. When the omnibus test was significant, pairwise Wilcoxon rank-sum tests were performed with Holm adjustment. All statistical tests were two-sided, and p < 0.05 was considered statistically significant. Analyses were performed in R using RStudio.

## RESULTS

### Study cohort

Twenty-four patients with PCABI admitted to the Vancouver General Hospital ICU between Oct 31, 2017 and Mar 14, 2023 underwent MRI and intraparenchymal PbtO_2_ monitoring and were included in the analysis. The patient cohort had a median age of 35 [29, 49] years. Most arrests occurred out of hospital (87.5%), with a median time to ROSC of 17.0 [13.5, 21.5] min, a median time from arrest to MRI of 3.0 [1.0, 8.0] days, and a median mean PbtO_2_ of 22.4 [14.8, 26.0] mmHg. Demographic, clinical, and physiological variables are summarized in Table 1 and Extended Data Table 1.

**Table 1.** Patient Characteristics.

| **Variable** | **Overall Cohort (N = 24<sup>1</sup>)** |
| --- | --- |
| **Age, years** | 34.5 (29.0, 49.0) |
| **Sex** |  |
| Female | 6 (25.0%) |
| Male | 18 (75.0%) |
| **BMI, kg/m<sup>2</sup>** | 25.4 (23.6, 26.1) |
| **Out-of-hospital cardiac arrest** | 21 (87.5%) |
| **Witnessed arrest** | 9 (37.5%) |
| **Shockable rhythm** | 5 (20.8%) |
| **Initial rhythm** |  |
| PEA | 16 (66.7%) |
| Asystole | 3 (12.5%) |
| VF | 5 (20.8%) |
| **Time to ROSC, min** | 17.0 (13.5, 21.5) |
| **Arrest etiology** |  |
| Anaphylaxis | 1 (4.2%) |
| Asthma | 1 (4.2%) |
| Cocaine | 2 (8.3%) |
| Electrocution | 1 (4.2%) |
| Hemorrhage | 2 (8.3%) |
| Hypoxia | 5 (20.8%) |
| Opioid overdose | 5 (20.8%) |
| Trauma | 7 (29.2%) |
| **Reactive pupils** | 23 (95.8%) |
| **Days from arrest to MRI** | 3.0 (1.0, 8.0) |
| **Mean PbtO<sub>2</sub>, mmHg** | 22.4 (14.8, 26.0) |
| **Survival** | 8 (33.3%) |
| **6-month CPC score** |  |
| CPC 1 | 2 (8.3%) |
| CPC 2 | 5 (20.8%) |
| CPC 3 | 1 (4.2%) |
| CPC 4 | 0 (0.0%) |
| CPC 5 | 16 (66.7%) |
| **Good neurological outcome (CPC 1-2)** | 7 (29.2%) |

<sup>1</sup>Median (Q1, Q3); n (%)

### PCABI burden is associated with lower PbtO_2_

Whole-brain lesion burden demonstrated an inverse correlation with mean PbtO_2_ (r = -0.49, p = 0.014), indicating greater MRI-defined injury burden is associated with lower PbtO_2_ (Figure 1B). Similar inverse associations were observed within both the left (r = -0.50, p = 0.012) and right (r = -0.48, p = 0.019) hemispheres (Figure 1C). Consistent with these findings, regional lesion burden demonstrated strong interhemispheric concordance across the most strongly affected white matter (r = 0.93-0.96, all p < 0.001) and grey matter regions (r = 0.86-0.97, all p < 0.001; Extended Data Figure 1). When stratified by tissue type, lesion burden within both grey matter (r = -0.48, p = 0.018) and white matter (r = -0.49, p = 0.016) was associated with lower PbtO_2_ (Figure 1D). Regional analyses demonstrated that the strongest associations between lesion burden and PbtO_2_ were localized to temporal and frontal brain regions (Figure 1E-L).

### MRI patterns of PCABI

PCA of regional MRI lesion burden data identified a dominant lesion-pattern axis, with PC1 and PC2 explaining 55.3% and 15.0% of total variance, respectively (70.3% cumulative variance; Figure 2A). PC1 scores were significantly associated with PbtO_2_ (r = 0.483, p = 0.0169; Figure 2B), indicating that variation in multiregional injury patterns was related to cerebral oxygenation. K-means clustering performed in PC1-PC2 space identified three distinct MRI injury patterns consisting of 13, 6, and 5 patients, respectively (Figure 2A). The lesion feature demonstrating the greatest separation across all three clusters was the left pars opercularis grey matter. Pairwise cluster comparisons identified the left entorhinal white matter as the most discriminative feature between Clusters 1 and 2 and between Clusters 2 and 3, whereas the left pericalcarine grey matter demonstrated the greatest separation between Clusters 1 and 3. Cluster assignment was highly stable across repeated k-means initializations, with identical clustering observed across all 100 random seeds. The clustering solution demonstrated clear separation and cohesion, with a mean silhouette score of 0.561 (Extended Data Figure 2). Cluster separation was also maintained when visualized across the first three principal components (Extended Data Figure 3).

### MRI-derived clusters demonstrate distinct regional injury patterns

Heatmap visualization of the top ranked cluster-discriminatory lesion features demonstrated clear separation between the three MRI-derived clusters (Figure 3; Extended Data Figure 4). Cluster 1 was characterized by uniformly low lesion burden across almost all grey and white matter regions, whereas Cluster 3 exhibited consistently elevated lesion burden across nearly all discriminatory features. Cluster 2 demonstrated an intermediate injury pattern with heterogeneous regional involvement. The lesion features contributing most strongly to separation of all three clusters included the pars opercularis, banks of the superior temporal sulcus, pericalcarine cortex, precuneus, superior parietal cortex, precentral gyrus, inferior parietal cortex, and putamen.

### Blood-based biomarkers across MRI-derived injury patterns

Arterial tau (p = 0.015) and GFAP (p = 0.009) varied across clusters, whereas NfL (p = 0.097) and UCH-L1 (p = 0.354) did not. Median [IQR] tau was higher in Cluster 2 (141.12 [105.97-144.89]) than Cluster 1 (3.71 [2.37-83.69]; p = 0.031). GFAP was lower in Cluster 3 (85.18 [84.74-87.23]) than Cluster 1 (372.50 [176.30-3192.40]) and Cluster 2 (1358.90 [1279.00-3192.40]; both p = 0.031; Figure 4B). No av_DIFF_ measures differed across clusters (Figure 4C).

### MRI-derived clusters represent distinct injury phenotypes

Whole-brain lesion burden differed significantly across MRI-derived clusters (p < 0.001; Figure 5A). Lesion burden increased progressively from Cluster 1 (10.16% [9.00-11.68]) to Cluster 2 (22.17% [17.83-25.12]) and Cluster 3 (38.40% [30.92- 41.45]). Admission arterial pH (p = 0.522), lactate (p = 0.801), serum creatinine (p = 0.612), and PaCO_2_ (p = 0.209) did not differ significantly across clusters (Figure 5B-E). Time to ROSC was not associated with whole-brain lesion burden (r = 0.30, p = 0.150; Figure 5F) or mean PbtO_2_ (r = -0.25, p = 0.247; Figure 5G). Mean PbtO_2_ did, however, decrease across MRI-derived clusters, with median values of 23.97 mmHg [IQR 20.13-25.33] in Cluster 1, 18.46 mmHg [14.49-22.29] in Cluster 2, and 12.49 mmHg [8.93-26.66] in Cluster 3 (Extended Data Table 1). Survival proportions were similar across MRI-derived clusters (Cluster 1: 4/13 [30.8%], Cluster 2: 2/6 [33.3%], Cluster 3: 2/5 [40.0%]), without a progressive increase in mortality with increasing lesion burden.

**Figure 5.**
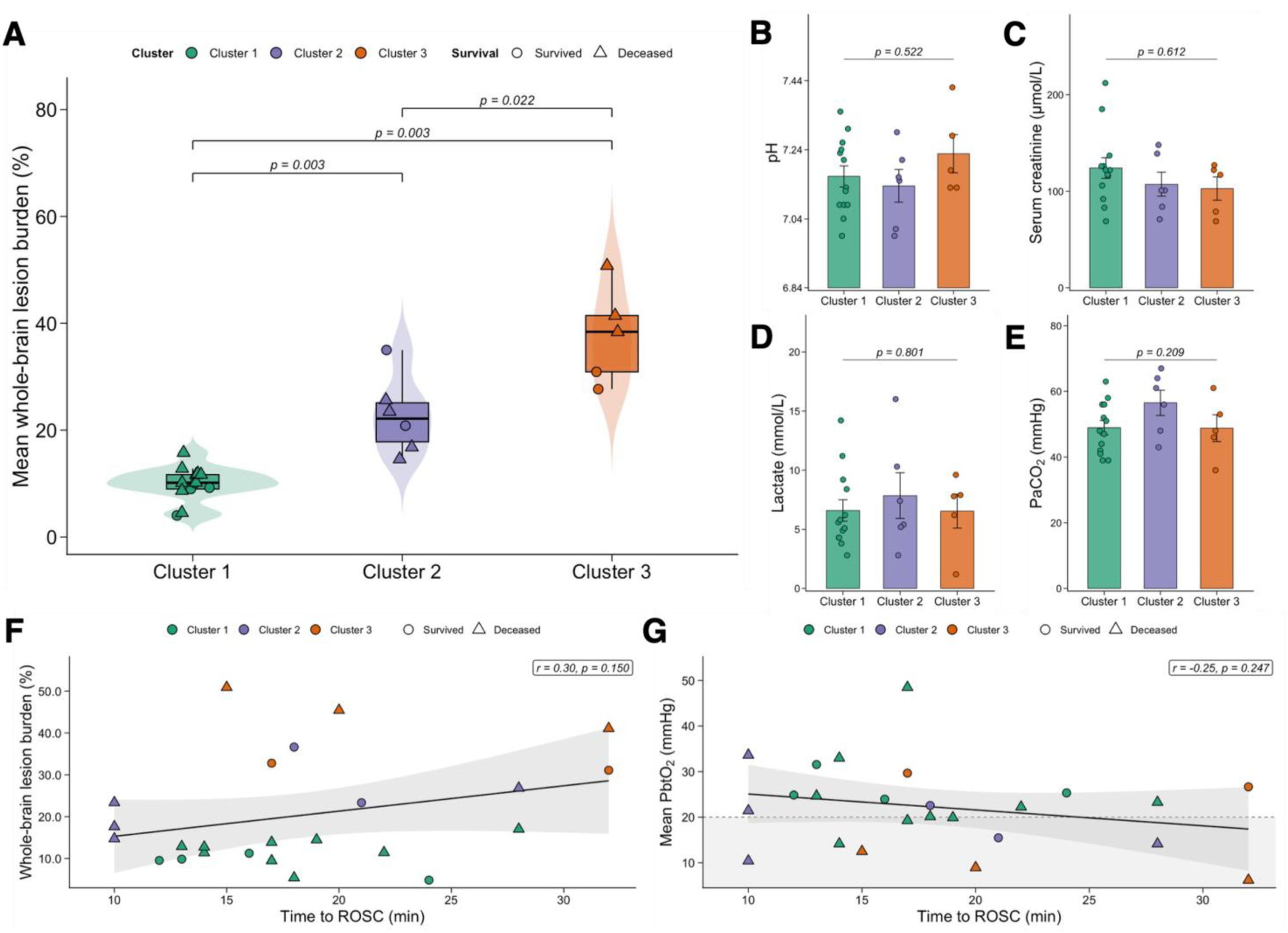
Whole-brain lesion burden, admission physiology, and time to return of spontaneous circulation across MRI-derived clusters. (A) Whole-brain ADC-defined lesion burden across MRI-derived clusters. Lesion burden differed across clusters by Kruskal-Wallis testing (p < 0.001), with significant Holm-adjusted pairwise differences between Cluster 1 and Cluster 2 (p = 0.003), Cluster 1 and Cluster 3 (p = 0.003), and Cluster 2 and Cluster 3 (p = 0.022). (B-E) Admission physiology across MRI-derived clusters, including admission arterial pH (p = 0.522), serum creatinine (p = 0.612), lactate (p = 0.801), and PaCO_2_ (p = 0.209). For panels B-E, p-values represent Kruskal-Wallis tests. Bars represent median values, error bars represent the interquartile range, and points represent individual patients. (F) Time to return of spontaneous circulation (ROSC) was not associated with whole-brain lesion burden (r = 0.30, p = 0.150). (G) Time to ROSC was not associated with mean brain tissue oxygen tension (PbtO_2_; r = -0.25, p = 0.247). For panels F-G, shaded bands represent 95% confidence intervals around the fitted regression lines. The dashed horizontal line in panel G indicates PbtO_2_ = 20 mmHg. Across panels, circles indicate survivors and triangles indicate deceased patients. In panel A, violin plots show the distribution of individual patient values; embedded boxplots show the median and interquartile range, with whiskers extending to 1.5 × interquartile range.

## DISCUSSION

In this retrospective analysis of patients with PCABI, MRI-based injury patterns were reflective of cerebrovascular physiology. Greater DWI/ADC lesion burden on clinically indicated MRI was associated with lower PbtO_2_ during the first 24 hours of neuromonitoring. This relationship was consistent across whole-brain, hemispheric, grey matter, white matter, and segment-level analyses. In parallel, multivariate analysis of anatomically resolved lesion features identified PCABI patterns that captured both a graded increase in global lesion burden and distinct spatial patterns of injury. These injury patterns were not associated with admission pH, lactate, creatinine, PaCO_2_, or time to ROSC alone, yet they were associated with PbtO_2_. Together, these findings indicate that physiologic phenotypes observed in patients with PCABI are associated with distinct underlying brain injury patterns.

The association between early PbtO_2_ and later MRI lesion burden provides an important physiological link between intraparenchymal neuromonitoring and spatiotemporal PCABI patterns. Prior studies have shown that brain hypoxia is common after cardiac arrest and can persist despite apparently acceptable systemic physiology.^21,24,27,55^ The present study extends this literature by demonstrating that lower PbtO_2_ corresponds to anatomically distinct DWI/ADC lesion burden in the subacute post-arrest period. Although the temporal separation between PbtO_2_ monitoring and MRI limits causal inference, the consistency of the association across multiple anatomical levels supports the interpretation that early tissue hypoxia and later diffusion-defined injury are potentially linked manifestations of PCABI severity and mechanism.

These findings fit directly within current models of PCABI as an evolving post-resuscitation syndrome rather than a fixed injury determined by ROSC duration.^1,2,4,13,14^ Indeed, time to ROSC was not associated with whole brain lesion burden, nor was time to ROSC different between injury patterns (Figure 5F; Supplementary Table S1). Moreover, these patterns were not explained by admission physiology (e.g., arterial lactate; Figure 5B-E). Arrest duration and systemic physiology remain central to PCABI, but they are incomplete proxies for cerebral injury biology.^17,56–63^ Time to ROSC does not capture no-flow duration, low-flow quality, hypoxemia, arrest etiology, resuscitation physiology, reperfusion injury, or secondary cerebral insults after ROSC.^1,2,4,17,56–59^ By applying atlas-based lesion quantification, this study shows that the oxygenation-injury relationship can be resolved across anatomical regions as well as tissue classes. This supports the view that spatial injury distribution contains clinically and biologically relevant information beyond total lesion volume and systemic arrest variables alone.^30,33–35,44–47^

The clustering analysis strengthens this interpretation by showing that PCABI can be separated into reproducible and distinct MRI-derived injury patterns.^18–20^ These injury patterns reflected both severity and spatial patterning: whole-brain lesion burden increased progressively across clusters, but cluster assignment was derived from multiregional lesion features rather than from global burden. This distinction is important because patients with similar total lesion burden may differ in anatomical injury distribution, tissue compartment involvement, and injury timing. Prior work has shown that diffusion abnormalities post-cardiac arrest are anatomically nonuniform and can evolve over time.^30,33–35,44–47^ The present findings build on that literature by identifying stable MRI-derived injury patterns.

The biomarker findings provide complementary evidence that MRI lesion patterns may differ in neuroglial injury biology, although this analysis is limited by small sample sizes and a single sampling timepoint. Arterial total tau and GFAP varied across MRI-derived clusters, whereas NfL and UCH-L1 did not. Total tau, NfL, GFAP, UCH-L1, and NSE have each been studied after cardiac arrest, with several demonstrating associations with neurological outcome.^21,64–73^ The lack of consistent biomarker differences may reflect a mismatch in biological scale. Circulating arterial biomarker concentrations and av_DIFF_ provide global measures of injury, whereas atlas-based MRI features capture spatially resolved injury patterns. Larger patient cohorts with serial arterial and jugular venous biomarker sampling are needed to define the temporal and mechanistic basis of these phenotypes.

To our knowledge, this is among the first studies to integrate anatomically resolved DWI/ADC lesion quantification with intraparenchymal PbtO_2_ monitoring and paired arterial-jugular venous biomarkers in patients with PCABI. Our atlas-based approach enabled lesion quantification across whole-brain, hemispheric, tissue-class, and segment-level features, building on prior semi-automated anatomical MRI quantification work in PCABI.^48^ Clustering was performed using MRI lesion features alone, reducing circularity between phenotype discovery and downstream physiological or biomarker interpretation. This design allowed us to test whether regional MRI lesion patterns correspond to PbtO_2_ and biomarker evidence of neuroglial injury, rather than treating MRI, PbtO_2_, and biomarkers as isolated prognosticators.

This was a retrospective, single-centre study with a limited sample size, and the association between MRI-derived injury patterns and cerebrovascular physiology requires external validation. PbtO_2_ was summarized over the first 24 hours of monitoring, whereas MRI was performed later, at a median of 3 days after arrest. This temporal separation supports the interpretation that early PbtO_2_ is associated with subsequent restricted diffusion-defined injury, but it limits causal inference. PbtO_2_ monitoring was unilateral and focal, typically within frontal white matter, and may not capture regional or interhemispheric variation in tissue oxygenation. Segment-level analyses therefore require confirmation in larger cohorts. Finally, this study outlines mechanistic phenotyping rather than prognostic model development, and the findings should not be used in isolation for neuroprognostication.

## CONCLUSION

Greater MRI-derived ischemic lesion burden was associated with decreased PbtO_2_ after cardiac arrest. Anatomically resolved DWI/ADC lesion features identified reproducible MRI-derived injury patterns that reflected both global injury severity and spatial lesion patterning. These findings demonstrate that integrating quantitative MRI lesion mapping with intraparenchymal neuromonitoring and blood-based biomarkers provides biologically informative characterization of PCABI rather than systemic admission variables, historical arrest characteristics, or global lesion burden alone. Future multicentre studies should validate these MRI-derived phenotypes, define their temporal evolution, and determine whether combining regional MRI lesion patterns with physiological and biomarker data improves mechanistic classification, risk stratification, or treatment selection after cardiac arrest.

## AUTHOR CONTRIBUTIONS

CAL: Conceptualization, Methodology, Software, Formal analysis, Investigation, Data curation, Validation, Visualization, Writing - original draft, Writing - review & editing. GA: Conceptualization, Methodology, Software, Formal analysis, Investigation, Data curation. WL: Resources, Data curation. SS: Investigation, Data curation. JC: Investigation, Data curation. CLW: Resources, Data curation. PAG: Resources, Data curation. DF: Resources, Data curation. DEG: Resources, Data curation. MSS: Resources, Data curation. WG: Conceptualization, Methodology, Resources, Data curation, Supervision. RLH: Conceptualization, Methodology, Formal analysis, Investigation, Data curation, Validation, Visualization, Resources, Supervision, Project administration, Funding acquisition, Writing - original draft, Writing - review & editing.

## DATA AVAILABILITY

The data are not publicly available due to patient privacy and institutional restrictions but may be available from the corresponding authors upon reasonable request and with appropriate institutional approvals.

