## Supplemental Material for "MRI-Derived Patterns of Post-Cardiac Arrest Brain Injury: Linking Anatomical Injury and Cerebrovascular Physiology"

**Affiliations**

### SUPPLEMENTAL METHODS

#### MRI lesion-data preparation and anatomical label processing

MRI lesion data were generated from the atlas-based DWI/ADC segmentation pipeline as described in the main methods. For downstream analysis, patient-level lesion data were retained by anatomical segment, hemisphere, tissue class, and lesion-mask volume. Pre-threshold and post-threshold segment volumes were arranged into paired columns for each patient, hemisphere, tissue class, and anatomical segment. Lesion burden was calculated as the percentage of the original segment volume meeting the ADC intensity-defined lesion criterion:

$$\text{lesion burden (\%)} = |\text{post-threshold volume} - \text{pre-threshold volume}| / \text{pre-threshold volume} \times 100$$

Segment labels were standardized prior to analysis. Left- and right-sided labels were identified using segment prefixes or text labels containing lh, rh, Left, or Right in aparc+aseg and wmparc atlases. After hemisphere assignment, hemisphere prefixes were removed from anatomical segment names to enable bilateral matching. Non-parenchymal and non-informative labels were excluded before downstream analyses. Excluded labels included CSF, third ventricle, fourth ventricle, inferior lateral ventricle, lateral ventricle, choroid plexus, vessel, white matter hypointensities, global cerebral white matter labels, and optic chiasm summary labels.

Each anatomical segment was assigned to grey matter or white matter using atlas-label patterns. White matter labels included the segment labels containing wm, cerebellar white matter, unsegmented white matter, and corpus callosum. Grey matter labels included cortical labels and subcortical grey matter structures, including cerebellar cortex, thalamus, caudate, putamen, pallidum, hippocampus, amygdala, accumbens area, and ventral diencephalon.

Standardized cortical and subcortical segment labels included the following: pericalcarine cortex, paracentral lobule, cuneus, caudal middle frontal cortex, precentral cortex, precuneus, superior parietal cortex, transverse temporal cortex, postcentral cortex, lateral occipital cortex, lingual gyrus, posterior cingulate, pars opercularis, banks of the superior temporal sulcus, isthmus cingulate, inferior parietal cortex, supramarginal gyrus, superior frontal cortex, pars orbitalis, pars triangularis, rostral middle frontal cortex, fusiform gyrus, inferior temporal cortex, superior temporal cortex, orbitofrontal regions, insula, middle temporal cortex, anterior cingulate regions, entorhinal cortex, parahippocampal cortex, frontal pole, thalamus, putamen, pallidum, hippocampus, amygdala, caudate nucleus, nucleus accumbens, ventral diencephalon, cerebellar cortex, cerebellar white matter, and unsegmented white matter.

This processing workflow was applied separately for whole-brain lesion burden, left-hemisphere lesion burden, right-hemisphere lesion burden, grey matter lesion burden, and white matter lesion burden.

#### Bilateral regional rankings for white matter and grey matter

For bilateral regional rankings, left- and right-hemisphere lesion percentages were averaged within each anatomical segment and tissue class before correlation testing with mean brain tissue oxygen tension ($PbtO_2$). Bilateral grey matter and white matter regions were ranked by Pearson correlation coefficient between regional lesion burden and mean $PbtO_2$. The top three bilateral grey matter regions and top three bilateral white matter regions were selected from these rankings for anatomical visualization and detailed correlation plots (Figure 1G-L).

#### Extended lesion data analysis

A segment-level lesion-distribution figure was generated using the 24-patient analytic cohort. Separate grey matter and white matter panels were generated using their respective assigned segments. Within each tissue class, lesion burden was displayed for the left hemisphere, right hemisphere, and left-right difference across corresponding anatomical segments (Extended Data Figure 1A-B).

Left-right agreement plots were generated for the top three grey matter regions and top three white matter regions identified from the bilateral $PbtO_2$ correlation rankings. For each selected segment, left-hemisphere lesion burden was plotted against right-hemisphere lesion burden. A unity line was shown, and Pearson correlation coefficients were calculated and displayed for each panel (Extended Data Figure 1C-H).

**Principal component analysis and MRI-derived clustering**
The PCA input matrix was generated from segment-level lesion percentages. Each MRI lesion feature was defined by the combination of anatomical segment, tissue class, and hemisphere. Midline or unpaired labels were retained (i.e. corpus callosum). The lesion dataset was converted into a patient-by-feature matrix. No data were imputed as there were no missing segment lesion values for any patient. Each lesion feature was centred and scaled across patients to generate z-scored feature values before PCA.

K-means clustering was performed using PC1 and PC2 scores only. The number of clusters was set to k = 3. Clustering was performed with 100 random starts and a fixed random seed of 12345678. After cluster assignment, clinical variables, mean $PbtO_2$, survival, 6-month Cerebral Performance Category (CPC), admission physiology, biomarker data, and outcome variables were joined to the clustered patient-level dataset for downstream characterization. None of these clinical, physiological, biomarker, or outcome variables were included in the PCA input matrix, k-means clustering, or final cluster assignment.

**Cluster validation and stability analyses**
Cluster validation was performed in PC1-PC2 space, matching the primary clustering space. For each candidate value of k, within-cluster sum of squares was calculated using k-means clustering with 100 random starts. Average silhouette width was calculated using Euclidean distances in PC1-PC2 space.

Repeated-seed stability was assessed across 100 seed values. For each candidate value of k, a reference clustering solution was generated using seed 12345678 and 100 random starts. K-means clustering was then repeated using seeds 1 through 100, with 100 random starts for each run. For the selected k = 3 solution, patient-level silhouette widths were calculated in PC1-PC2 space using Euclidean distances, and the average silhouette width was calculated across all patients. Patient-level co-clustering stability was also calculated for the selected k = 3 solution. For each repeated-seed run, every pair of patients was coded as co-clustered if both patients were assigned to the same cluster. A co-clustering frequency matrix was then calculated as the proportion of repeated runs in which each patient pair was assigned to the same cluster.

**Robustness to additional principal components**
The primary k = 3 clustering solution based on PC1-2 was compared with a higher-dimensional clustering solution using PC1 through PC12. K-means clustering was repeated using PC1-12 with k = 3, 100 random starts, and seed 12345678. Agreement between the PC1-2 solution and the PC1-12 solution was quantified using the adjusted Rand index (ARI). The ARI between the PC1-PC2 and PC1-PC12 clustering solutions was 1.0, indicating perfect agreement between the two solutions; therefore, inclusion of additional principal components did not alter patient cluster assignments. This analysis was used only as a cluster-assignment robustness check and was not used for downstream analyses.

**Selection of discriminatory MRI lesion features**
After cluster-assignment, MRI lesion features were ranked by their ability to distinguish MRI-derived clusters. For each lesion feature, Kruskal-Wallis testing was used to compare lesion burden across all three clusters. Pairwise Wilcoxon rank-sum tests were then also performed for Cluster 1 versus Cluster 2,

Cluster 1 versus Cluster 3, and Cluster 2 versus Cluster 3, with p-values adjusted using the Holm method. For the overall three-cluster feature panel, the top feature was selected from the Kruskal-Wallis ranking. For pairwise feature panels, the most discriminatory feature for each cluster comparison was selected as the feature with the smallest adjusted pairwise p-value for that comparison.

**Heatmap generation**
Heatmaps were generated from standardized lesion-feature values. For heatmap input, the patient-by-feature lesion matrix was row-scaled so that each MRI lesion feature was displayed as a z-score across patients. Patients were ordered by MRI-derived cluster assignment and, within each cluster, by mean $PbtO_2$.

For the main heatmap (Figure 3), features were selected separately within four anatomical classes: left grey matter, right grey matter, left white matter, and right white matter. Within each anatomical class, the top 10 features were selected. For each feature, median lesion burden was calculated within Cluster 1, Cluster 2, and Cluster 3. Pairwise median differences were calculated for Cluster 1 versus Cluster 2, Cluster 2 versus Cluster 3, and Cluster 1 versus Cluster 3. A balanced discrimination score was then calculated as:

$$\text{Balanced discrimination score} = \min(|\text{Cluster 1} - \text{Cluster 2}|, |\text{Cluster 2} - \text{Cluster 3}|) + 0.25 \times |\text{Cluster 1} - \text{Cluster 3}|$$

This score was used only to select representative features for heatmap visualization and was not used for PCA, k-means clustering, or statistical inference. The final selected-feature heatmap contained 40 features in total, 10 from each of the four tissue classes included. Extended Data Figure 4 displays the complete set of anatomical lesion features included in the clustering analysis. The same z-scored patient-by-feature matrix and patient ordering were used for this extended heatmap that contains all MRI segments.

**Software**
Analyses were performed in R. The fixed random seed used for reproducible analyses was 12345678. Packages used across the analysis workflow included tidyverse, ggpubr, kableExtra, nlme, emmeans, multcomp, broom, broom.mixed, factoextra, cluster, ggrepel, readxl, janitor, gtsummary, gt, cocor, patchwork, scales, mclust, rstatix, pheatmap, and plotly.

**SUPPLEMENTAL TABLES**

Extended Data Table 1. Clinical, physiological, and MRI characteristics across MRI-derived injury patterns.

| **Variable** | **Cluster 1 (n = 13)** | **Cluster 2 (n = 6)** | **Cluster 3 (n = 5)** |
|---|---|---|---|
| Whole brain lesion (%) | 9.99 ± 0.88 | 22.74 ± 2.97 | 37.85 ± 4.07 |
| Left hemisphere lesion (%) | 9.27 ± 0.91 | 22.41 ± 2.61 | 38.24 ± 4.11 |
| Right hemisphere lesion (%) | 10.87 ± 0.97 | 23.71 ± 3.21 | 39.15 ± 4.45 |
| White matter lesion (%) | 9.88 ± 1.02 | 25.54 ± 4.45 | 42.39 ± 4.12 |
| Grey matter lesion (%) | 9.98 ± 0.79 | 20.21 ± 2.03 | 34.00 ± 4.31 |
| Mean $PbtO_2$ (mmHg) | 25.45 ± 2.36 | 19.61 ± 3.37 | 16.78 ± 4.78 |
| Admission arterial pH | 7.16 ± 0.03 | 7.13 ± 0.05 | 7.23 ± 0.06 |
| Admission lactate (mmol/L) | 6.60 ± 0.91 | 7.85 ± 1.92 | 6.54 ± 1.44 |
| Admission $PaCO_2$ (mmHg) | 48.92 ± 2.16 | 56.50 ± 3.84 | 48.80 ± 4.12 |
| Admission creatinine (μmol/L) | 124.08 ± 10.69 | 107.33 ± 12.38 | 102.80 ± 11.97 |
| Time to ROSC (min) | 17.46 ± 1.32 | 16.17 ± 3.06 | 23.20 ± 3.68 |

Values are mean ± SEM.

### SUPPLEMENTAL FIGURES

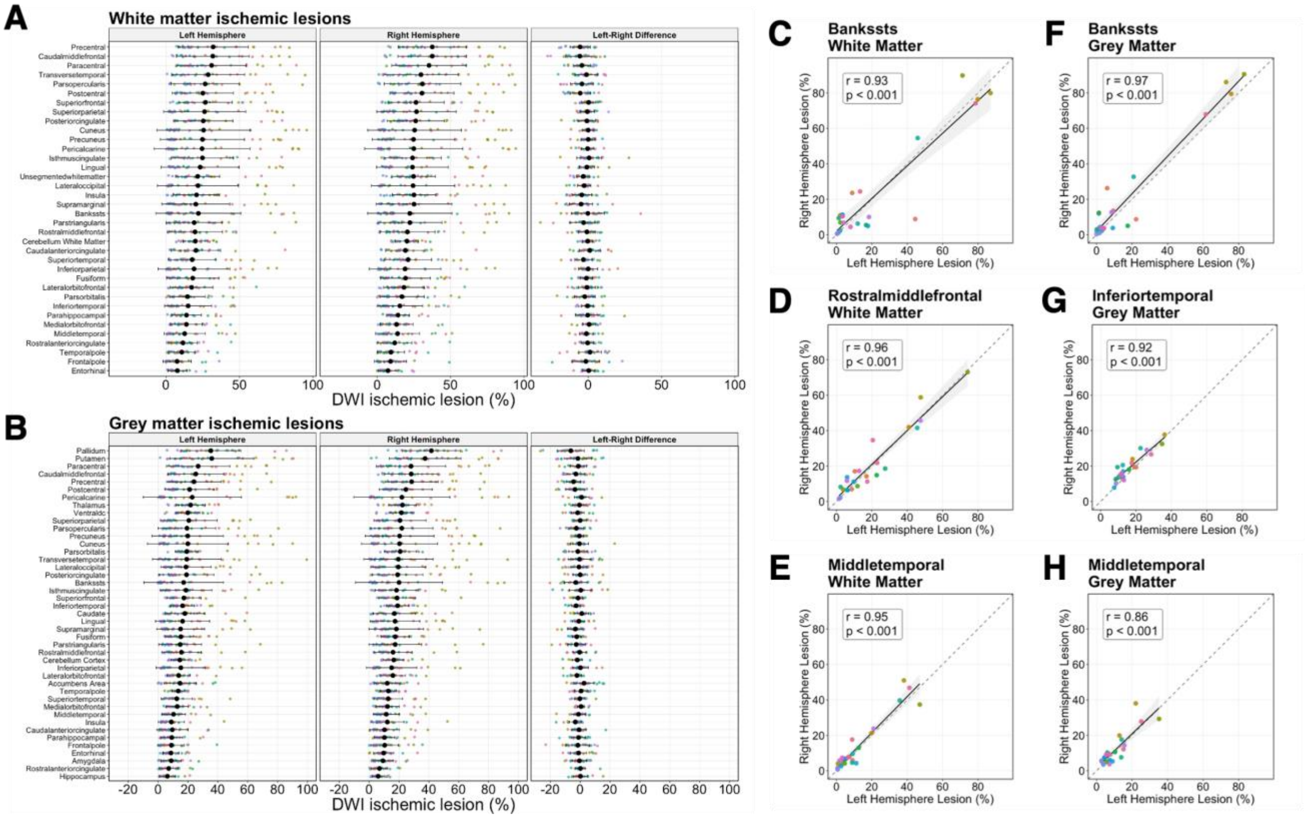


**Extended Data Figure 1. Distribution and hemispheric agreement of grey and white matter ischemic lesion burden.** (A) White matter ischemic lesion burden (%) across atlas-defined segments, shown for the left hemisphere, right hemisphere, and the left–right difference across corresponding segments. (B) Grey matter ischemic lesion burden (%) across atlas-defined segments. (C-E) Agreement between left and right hemisphere lesion burden (%) for the top three white matter segments, demonstrating strong interhemispheric concordance: (C) $r = 0.93$, $p < 0.001$; (D) $r = 0.96$, $p < 0.001$; (E) $r = 0.95$, $p < 0.001$. (F-H) Agreement between left and right hemisphere lesion burden (%) for the top three grey matter segments, also demonstrating strong interhemispheric concordance: (F) $r = 0.97$, $p < 0.001$; (G) $r = 0.92$, $p < 0.001$; (H) $r = 0.86$, $p < 0.001$. Lesion burden was derived from atlas-based segmentation and expressed as percentage of regional volume.

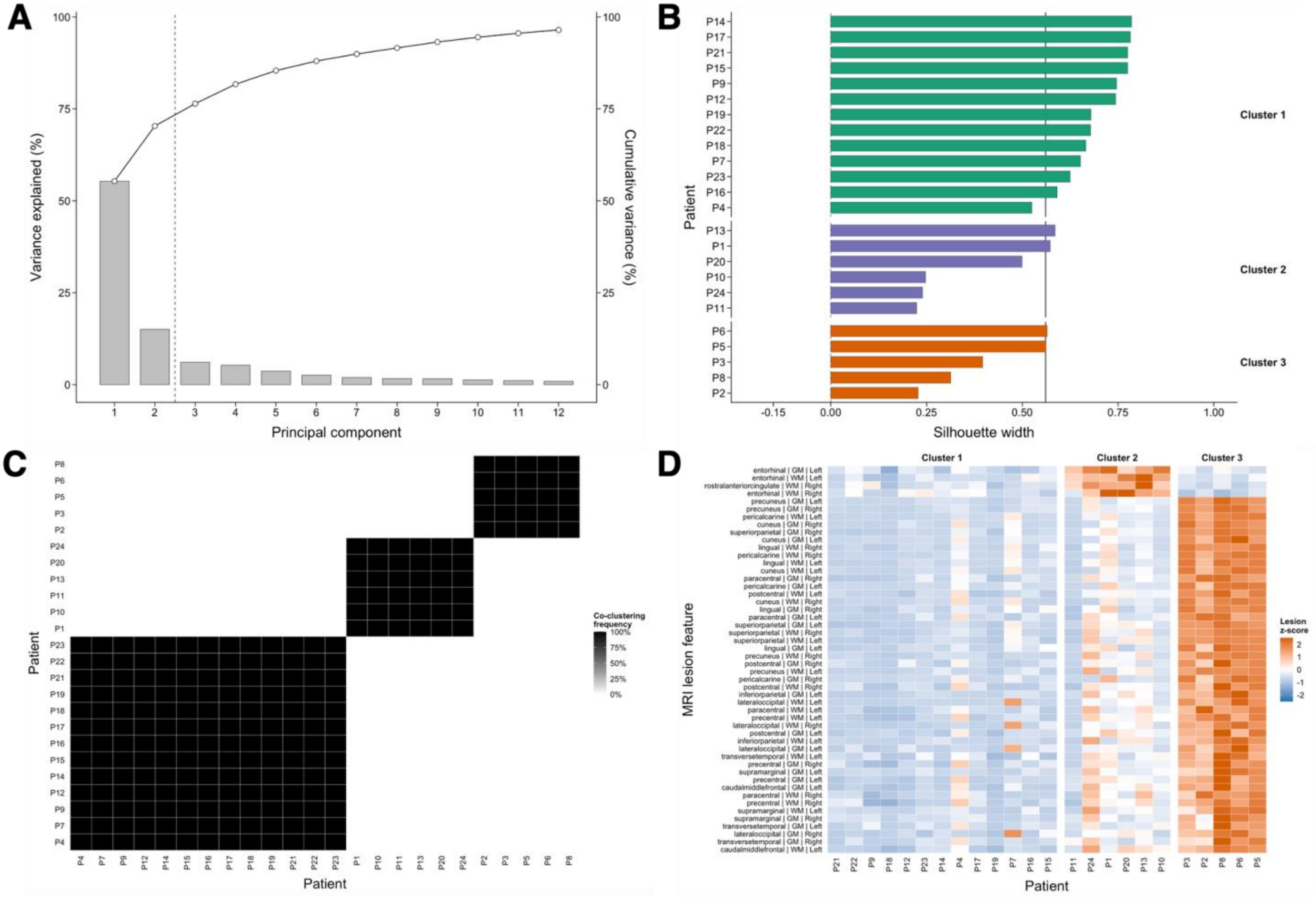


**Extended Data Figure 2. Validation of MRI-derived clustering in PC1-PC2 space.** (A) Principal component analysis (PCA) variance explained across components. The first two principal components (PC1-PC2), which were used for clustering, explain 70.3% of the total variance in the dataset. (B) Patient-level silhouette widths in PC1-PC2 space stratified by cluster, indicating generally positive within-cluster cohesion and separation between clusters. (C) Repeated-seed co-clustering stability analysis in PC1-PC2 space across 100 repeated-seed runs, demonstrating high reproducibility of cluster assignments. (D) Top MRI lesion-pattern features ranked by between-cluster separation, derived from z-scored lesion feature matrices, highlighting the dominant imaging signatures differentiating clusters. Clustering was performed using k-means on PC1-PC2 scores only (no additional PCs included in the clustering model). All analyses are descriptive and intended to assess stability and interpretability of the MRI-derived clustering solution (n = 24).

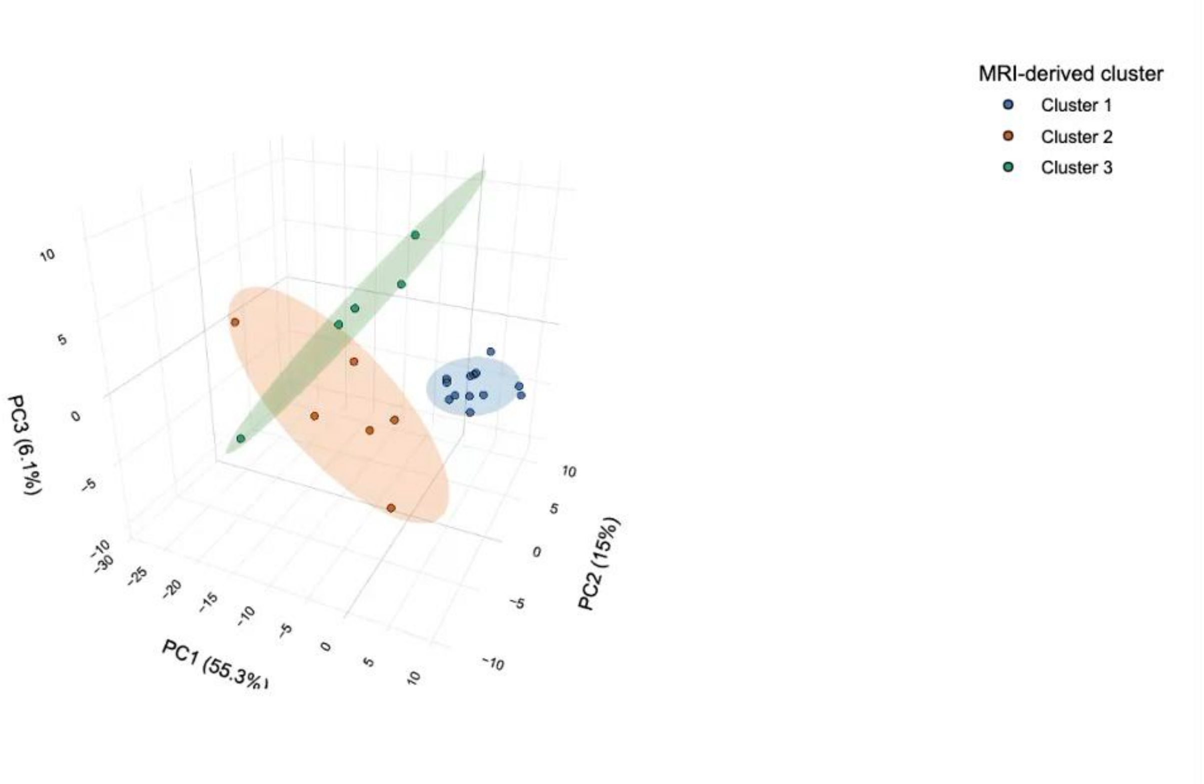


**Extended Data Figure 3. Three-dimensional PCA visualization of cluster structure.** Three-dimensional PCA projection (PC1-PC3) of MRI lesion clusters. The first three components explain 76.4% of total variance. Patients are coloured by cluster assignment derived from k-means clustering in PC1-PC2 space. Clusters remain visually separable in higher-dimensional PCA space, supporting robustness of the observed grouping structure.

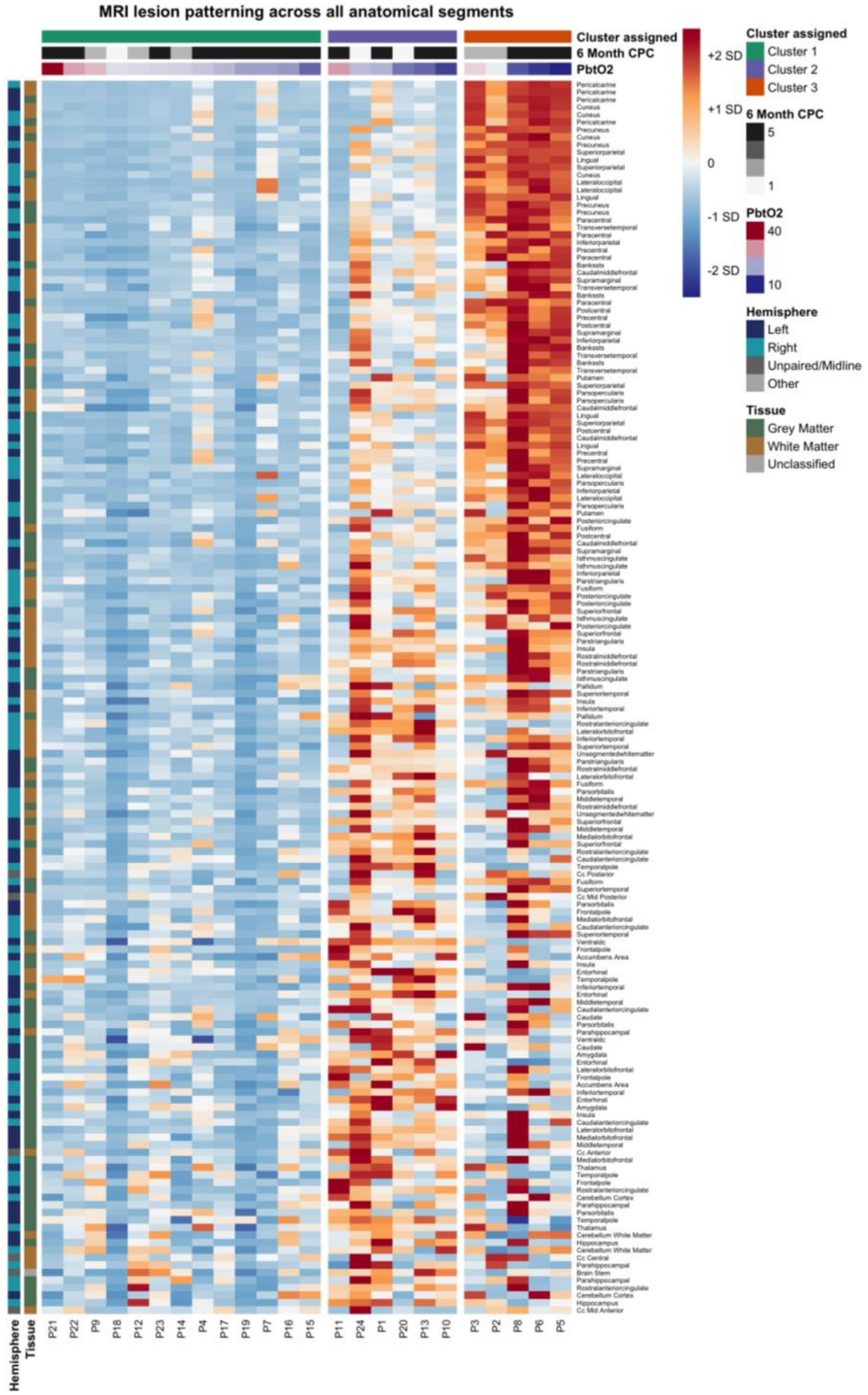


**Extended Data Figure 4. MRI lesion pattern heatmap across all anatomical segments.** Heatmap of standardized MRI lesion feature z-scores across patients grouped by MRI-derived cluster assignment. Columns represent individual patients and rows represent all anatomical lesion segments included in the clustering analysis, spanning white matter and grey matter regions in the left and right hemispheres. Patients are ordered by cluster assignment and, within each cluster, by brain tissue oxygenation ($PbtO_2$). Annotation tracks indicate cluster membership, 6-month Cerebral Performance Category (CPC), and $PbtO_2$. This extended figure displays the complete set of lesion segments used for clustering.
